# Serosurvey of SARS-COV-2 at a large public university

**DOI:** 10.1101/2023.02.05.23285494

**Authors:** Ching-Wen Hou, Stacy Williams, Kylee Taylor, Veronica Boyle, Bradley Bobbett, Joseph Kouvetakis, Keana Nguyen, Aaron McDonald, Valerie Harris, Benjamin Nussle, Phillip Scharf, Megan Jehn, Timothy Lant, Mitch Magee, Yunro Chung, Joshua Labaer, Vel Murugan

## Abstract

Arizona State University (ASU) is one the largest universities in the United States, with more than 79,000 students attending in-person classes. We conducted a seroprevalence study from September 13-17, 2021 to estimate the number of people in our community with SARS-CoV-2-specific antibodies due to previous exposure to SARS-CoV-2 and/or vaccination. Participants provided their age, gender, race, status (student or employee), and general COVID-19 health-related information like previous exposure and vaccination status. The seroprevalence of the anti-receptor binding domain (RBD) antibody was 90% by a lateral flow assay and 88% by a semi-quantitative chemiluminescent immunoassay. The seroprevalence for anti-nucleocapsid (NC) was 20%. In addition, individuals with previous natural COVID infection plus vaccination had higher anti-RBD antibody levels compared to those who had vaccination only or infection only. Individuals who had a breakthrough infection had the highest anti-RBD antibody levels.

Accurate estimates of the cumulative incidence of SARS-CoV-2 infection can inform the development of university risk mitigation protocols such as encouraging booster shots, extending mask mandates, or reverting to online classes. It could help us to have clear guidance to take action at the first sign of the next surge as well, especially since there is a surge of COVID subvariant infections.

## Introduction

The COVID-19 pandemic has been a major challenge worldwide. COVID-19 is caused by a novel Betacoronavirus (SARS-CoV-2) and was first reported from Wuhan, China, on 8 December 2019. The World Health Organization (WHO) declared it a pandemic on 11 March 2020 ^1,2^. The United States had recorded more than 101 million cases and 1,091,000 deaths by January 11, 2023. Since the end of 2019, communities around the world have had to fight against outbreaks, including physical distancing, staying at home, avoiding groups indoors, wearing masks, frequent testing, and contact tracing, etc ^3^. Although the intensity of these measures has recently abated partially, activities have not fully returned to the pre-pandemic routine and there are still an estimated 2500 COVID-19 deaths weekly^4^. Scientists have developed rapid diagnostics tests ^5-7^ and many effective vaccines ^8-10^ that have reduced morbidity and mortality considerably. Throughout the pandemic, university life has represented a unique challenge because universities tried to maximize safe in-person learning opportunities and maintain safe school operations by implementing effective practices.

ASU shifted to online classes on March 16, 2020 while a team of researchers at the Biodesign Institute set up a clinical testing laboratory. At that time, Arizona was a worldwide COVID-19 hotspot. ASU students and employees were encouraged to do COVID test frequently at no charge. After a few months of monitoring COVID, ASU switched to hybrid classes in August 2020. However, COVID cases began surging in late November 2020, resulting in the implementation of a fully remote learning model in December 2020. On January 11, 2021, ASU switched back to hybrid learning model until Fall semester of 2021. During these months, COVID testing, and vaccines were available to all students and employees at ASU. The ASU community followed CDC guidelines by offering frequent qPCR saliva testing, rigorous contact tracing, and strong support during isolation. This allowed the safe return to a fully in-person class in the fall of 2021 (Figure 1).

**Figure 1.**
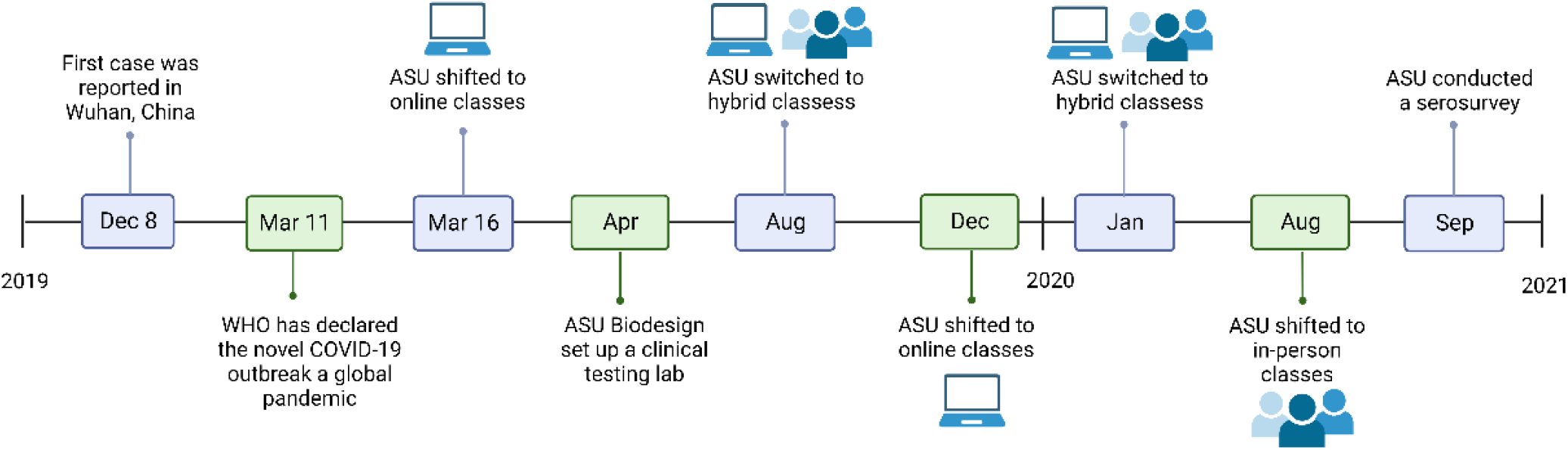
The timeline between the outbreak of the COVID-19 pandemic and serosurvey at ASU. In response to COVID, ASU rotated from in-person to remote to hybrid learning several times during the pandemic depending on the prevalence of infection in the community.

A research project at Davidson College in North Carolina reported that almost 6000 four-year colleges and universities provided combinations of online and in-person classes, another 446 had “primarily in-person” courses, and 45 operated “fully in-person” during 2020^11^. Also, a survey was conducted by New America and Global Strategy Group with 1,002 college students nationwide from April 29 through May 13, 2021. In the survey, 62% of students claimed that their schools would provide combinations of online and in-person classes, 14% claimed that their schools will offer online classes only, and only 12% would provide fully in-person classes in the fall of 2021^12^. Arizona State University was among the 12% operating “fully in-person” and one of the country’s largest universities, with over 79,000 students who had returned to campus for in-person classes in the fall of 2021. ASU wanted to evaluate the success of the COVID-19 management strategy by monitoring SARS-CoV-2 seroprevalence to estimate immunity from prior infection, vaccination, or both. Thus, ASU conducted a serosurvey to collect self-reported experiences and to determine the number of people in our community with various SARS-CoV-2-specific antibodies. The university had anonymized information about the prevalence of positive qPCR tests in its community, but at the time of this study, it lacked information about the level of immunity and possible viral exposure rate. This study would help inform on deciding on safety protocols, vaccination recommendations, masking recommendations mandates, and online vs in-person classes. At the time of this survey, September 2021, the SARS-CoV-2 subvariant Delta, a highly contagious variant, accounted for 65% of all cases in Arizona ^4^.

By assessing humoral immunity, seroprevalence studies estimate the percentage of a specific population who have been previously infected with a pathogen. Many seroprevalence studies of SARS-CoV-2 have been reported. Arnaud et al. showed that neutralizing and anti-RBD antibodies persisted for at least 6 months after a mild COVID infection from hospital workers^13^. A similar result was also demonstrated by Baker et al, who found that the antibodies against SARS-CoV-2 produced by health care workers or patients who have mild COVID infection were stable for up to six months and helped prevent recurrent infections ^14^. Another group investigated anti-NC antibody levels in severe and mild patients at hospitals. The data indicated that anti-NC levels started to decline after 2 months after post PCR and antibody levels were lower in patients with mild compared to severe illness ^15^. However, the seroprevalence studies at the educational institutions/ communities, where students and employees are in close contact on daily basis, are very limited.

SARS-CoV-2 induces antibodies with IgM, IgA, and IgG isotypes against spike protein, RBD of the spike protein, and NC protein. The antibodies produced by COVID vaccination (Pfizer, Moderna, J&J, AstraZeneca, and Covishield) are IgM, IgA, and IgG isotypes against spike protein, specifically the RBD of the spike protein. Whereas anti-spike antibodies do not distinguish between vaccination and infection, anti-NC positivity generally implies a previous infection; however, participants who received COVID vaccines from Sinopharm, Sinovac, and Covaxin, which contain inactivated SARS-CoV-2 viruses, and who attend an international university like ASU, may also have anti-NC antibodies from vaccination. By combining the self-reported vaccine and infection history with documented antibodies to SARS-CoV2 antigens, we estimated the number of: a) individuals with detectable anti-spike antibodies; b) individuals with likely previous SARS-CoV-2 exposure, even if they did not report COVID-19 symptoms; and c) individuals with no detectable antibodies after vaccination or previous infection.

This study took volunteers over five days (9/13-9/17 of 2021) and 1064 serum samples and matching survey results were analyzed. The serological tests were done either at the ASU Biodesign Clinical Testing Laboratory (ABCTL) or the Center for Personalized Diagnostics (CPD), using multiple EUA-approved assays targeting antibodies against the NC protein and spike proteins. The sera for all 1064 participants were tested using a lateral flow assay (Sienna-Clarity COVIBLOCK COVID-19 IgG/IgM Rapid Test) and a chemiluminescent assay (Beckman Access SARS-CoV-2 IgG II and IgM) to detect anti-RBD IgG and IgM. In addition, sera were also tested using a lateral flow assay (Megna) and ELISA assay (BioRad-Platelia SARS-CoV-2 Total Ab Assay) to detect anti-NC antibodies. The manufacturer-reported sensitivity and specificity are: a) 96.15% and 100% for Clarity; b) 98.9% and 100% for Beckman; c) 100% and 98.86% for Bio-Rad; and d) 95% and 99.3% for Megna. Participants reported that they were 92% vaccinated, which agreed with a 90% seroprevalence for anti-spike antibodies. Seroprevalence for anti-NC was found to be almost 19.5%, which included 11% from the participants who self-reported to have had a previous COVID infection and 8.5% from the participants reporting no known history of infection (Table 4).

## Methods

### Participants

We employed a two-stage sampling strategy. First, a random sample of current students were invited to participate in the serosurvey through email invitation. To increase the representativeness of the sample, targeted recruitment was made via social media advertising, as well as in-person recruitment from selected areas of the campus. Responses are time-stamped to allow for analysis according to the date of completion.

### Ethical approval

The study was approved by ASU’s institutional Review Board (IRB)(STUDY00014505). All participants are 18+ years old and consented to participating in the study and were willing to provide their samples for the research.

### Survey Instruments

Demographics, COVID-19 vaccination, testing history, and COVID symptoms were self-reported by questionnaire.

### Blood sample collection

The blood samples were collected by phlebotomists at ASU with serum tubes (Cat # 37988 from BD) and were placed into a cooler within 4 hours and transported within 6 hours of collection to the clinical testing laboratory at ASU. Samples were centrifuged at 1300 g for 20 minutes to separate serum.

### Serology testing

The main serological detection methods were all approved by Emergency Use Authorization from the US Food and Drug Administration for marketing are the chemiluminescence immunoassay (CLIA), enzyme-linked immunosorbent assay (ELISA), immunofluorescence assay (IFA), and lateral flow immunoassay (LFA) ^16,17^. In this survey, samples were tested for antibodies against the RBD domain of the Spike protein using Access SARS-CoV-2 chemiluminescent IgG II and IgM assay (Beckman coulter) and Sienna-Clarity COVIBLOCK COVID-19 IgG/IgM Rapid lateral flow assay (Sienna-Clarity) to estimate vaccine-induced SARS-CoV-2 seroprevalence. Samples also were tested for antibodies against table NC protein using Platelia SARS-CoV-2 Total Ab ELISA Assay and rapid COVID-19 IgM/IgG Combo lateral flow test kit (Megna Health Inc.) to estimate infection-induced SARS-Cov-2 seroprevalence (Supplementary Table 1).

Access SARS-CoV-2 chemiluminescent IgG II and IgM assays from Beckman coulter were performed in this study to determine IgG and IgM antibody level of SARS-coV-2 RBD protein according to the manufacturer’s instructions ^18^. 5 different concentrations of calibrators and two different concentrations of controls were provided by the manufacture to ensure reagent integrity and proper assay performance before analyzing samples. The result is compared to the cut-off value defined during the calibration of the instrument.

Sienna-Clarity COVIBLOCK COVID-19 IgG/IgM Rapid lateral flow assay is to detect IgG and/or IgM isotypes specific to the RBD portion of the S1 protein. 10 μL of serum and 2 drops of buffer were added, and test results were read after 10 min by the laboratory technician and the kits were photographed for a second independent reading by another laboratory technician.

Platelia SARS-CoV-2 total Ab ELISA assay from Bio-Rad is a qualitative diagnostic test. It is the detection of total antibodies (IgM/IgA/IgG) against SARS-CoV-2 NC. The result was interpreted based on the manufacturer’s recommendations: < 0.8, negative; between > 0.8 and < 1.0, equivocal; ≥1.0, positive.

COVID-19 IgM/IgG Combo lateral flow test kit from Megna Health Inc is to detect IgG and/or IgM isotypes specific to NC protein. 2 μL of serum and 2 drops of buffer were added and test results were read after 15 min by the laboratory technician and the kits were photographed for a second independent reading by another laboratory technician.

Meso scale discovery (MSD) coronavirus panel from Meso Scale Diagnostics is a multiplexed immunoassay to measure the IgG antibody response to SARS-CoV-2. A 96-well MSD plate has different antigens in each well. A calibration curve was created by using a reference standard with 4-fold serial dilution steps and a zero-calibrator blank for quantitation. Three levels of controls were also included in the assay to ensure the accuracy of the performance. First, the plate was blocked with Blocker A solution for 30 minutes at RT. The plate was washed 3 times with 150 μL/well of MSD wash buffer and then 50 μL of calibrator, controls, and diluted samples were dispensed into the plate and incubated with shaking for 2 hours at RT. After incubation and 3 × 150 μL/well washes with MSD wash buffer, detection antibody was added and then incubated with shaking for 1 hour. After detection antibody, the plate was washed with wash buffer following which reader buffer B was added and the plate reads using the MESO QuickPlex SQ 120 instrument.

## Results

### Demographic

Overall, this survey included 1064 participants from the four different campuses of ASU. A total of 480 students were randomly selected to receive invitation emails, which led to 250 subjects (28% of the final student participants); the remaining participants responded to university wide advertising, learned of the survey by word of mouth or personally observed the collections and volunteered. The participants provided saliva samples for a qPCR diagnostic test and also donated blood. Of the 1064 participants, 893 (83.9%) subjects were students, 79 (7.4%) were employees, and 92 (8.6%) subjects did not provide the information of the occupation status. 556 participants (52.3%) were female, and 467 participants (43.9%) were male. 762 participants (71.6%) were in the age group of 18-25 years, 190 (17.9%) were aged 26-40 years, 81 (7.6%) were aged 41-65 years, 31 (2.9%) were not reported. The demographic characteristics of the three different groups are presented in Table 1.

**Table 1.** Demographics of our serosurvey participants

|  |  | **Students**<br>**(n=893)** | **Employees**<br>**(n=79)** | **Randomly**<br>**Selected\***<br>**(n=250)** | **Total**<br>**Participants**<br>**(n=1064)** |
| --- | --- | --- | --- | --- | --- |
| **Gender** | Female | 444 (49.7%) | 51 (64.6%) | 142 (56.8%) | 556 (52.3%) |
|  | Male | 409 (45.8%) | 28 (35.4%) | 101 (40.4%) | 467 (43.9%) |
|  | Other | 10 (1.1%) | NA | 2 (0.8%) | 11 (1.0%) |
|  | Not Reported | 30 (3.4%) | NA | 5 (2%) | 30 (2.8%) |
| **Age** | 18-25 | 723 (81%) | 4 (5.1%) | 183 (73.2%) | 762 (71.6%) |
|  | 26-40 | 120 (13.4%) | 31 (39.2%) | 50 (20%) | 190 (17.9%) |
|  | 41-65 | 19 (2.1%) | 44 (55.7%) | 12 (4.8%) | 81 (7.6%) |
|  | Not Reported | 31 (3.5%) | NA | 5 (2%) | 31 (2.9%) |
| **Race** | White | 410 (45.9%) | 62 (78.5%) | 121 (48.4%) | 528 (49.6%) |
|  | Asian | 270 (30.2%) | 6 (7.6%) | 66 (26.4%) | 292 (27.4%) |
|  | Mixed | 39 (4.4%) | 5 (6.3%) | 13 (5.2%) | 46 (4.3%) |
|  | Black | 24 (2.7%) | 1 (1.3%) | 7 (2.8%) | 27 (2.5%) |
|  | Native | 13 (1.5%) | NA | 6 (2.4%) | 14 (1.3%) |
|  | Other | 99 (11.1%) | 4 (5.1%) | 31 (12.4%) | 117 (11%) |
|  | Prefer not to say | 7 (0.8%) | 1 (1.3%) | 1 (0.4%) | 9 (0.9%) |
|  | Not Reported | 31 (3.5%) | NA | 5 (2%) | 31 (2.9%) |
| **Vaccination Status** | Yes | 822 (92.1%) | 70 (88.6%) | 239 (95.6%) | 978 (91.9%) |
|  | No | 67 (7.5%) | 9 (11.4%) | 11 (4.4%) | 82 (7.7%) |
|  | Not Reported | 4 (0.5%) | NA | NA | 4 (0.4%) |
| **Vaccine Source** | Pfizer | 424 (47.5%) | 32 (40.5%) | 137 (54.8%) | 510 (47.9%) |
|  | Moderna | 248 (27.8%) | 35 (44.3%) | 70 (28%) | 309 (29.0%) |
|  | Janssen | 86 (9.6%) | 2 (2.5%) | 18 (7.2%) | 94 (8.8%) |
|  | AstraZeneca | 46 (5.2%) | NA | 9 (3.6%) | 46 (4.3%) |
|  | Covaxin | 9 (1.0%) | NA | NA | 9 (0.9%) |
|  | Sinopharm | 2 (0.2%) | NA | 2 (0.8%) | 2 (0.2%) |
|  | Sinovac | 1 (0.1%) | NA | 1 (0.4%) | 1 (0.1%) |
|  | Not Reported | 77 (8.6%) | 1 (1.3%) | 13 (5.2%) | 93 (8.7%) |
| **previous self-reported Covid infection** | Yes | 174 (19.5%) | 12 (15.2%) | 32 (12.8%) | 205 (19.3%) |
|  | No | 717 (80.3%) | 67 (84.8%) | 218 (87.2%) | 857 (80.6%) |
|  | Not Reported | 2 (0.2%) | NA | NA | 2 (0.2%) |

\*Randomly selected from enrolled students and invited by email

### Self-reported COVID-19 infection and vaccine status

Asymptomatic carriers can be a potential source of infection outbreaks in the community. We therefore evaluated how many participants had active COVID without reporting symptoms on the day when they donated samples. We found the prevalence of PCR positivity in asymptomatic students and employees in the university community was 0.4% (n=4/1064) on the day of sample collection. Among the 1064 participants, nearly 20% (19.3%, n=205/1064) reported testing positive for COVID-19 test in the past, whereas 80.6% reported no history of a positive test (Table 3).

More than 90% (91.9%, n=978/1064) of participants reported at least 1 dose of vaccine, whereas 7.7% of participants reported never receiving vaccine. Most participants received Pfizer (47.9%, n=510/1064) and Moderna (29.0%, n=309/1064) (Table 1). There was no significant difference in vaccine rate, nor reported history of COVID across age groups; however, we noticed that the lowest COVID rate among the 26-40 years group (14.2%, n=27/190) had the highest vaccine rate (93.2%, n=177/190) (Table 2).

**Table 2.**
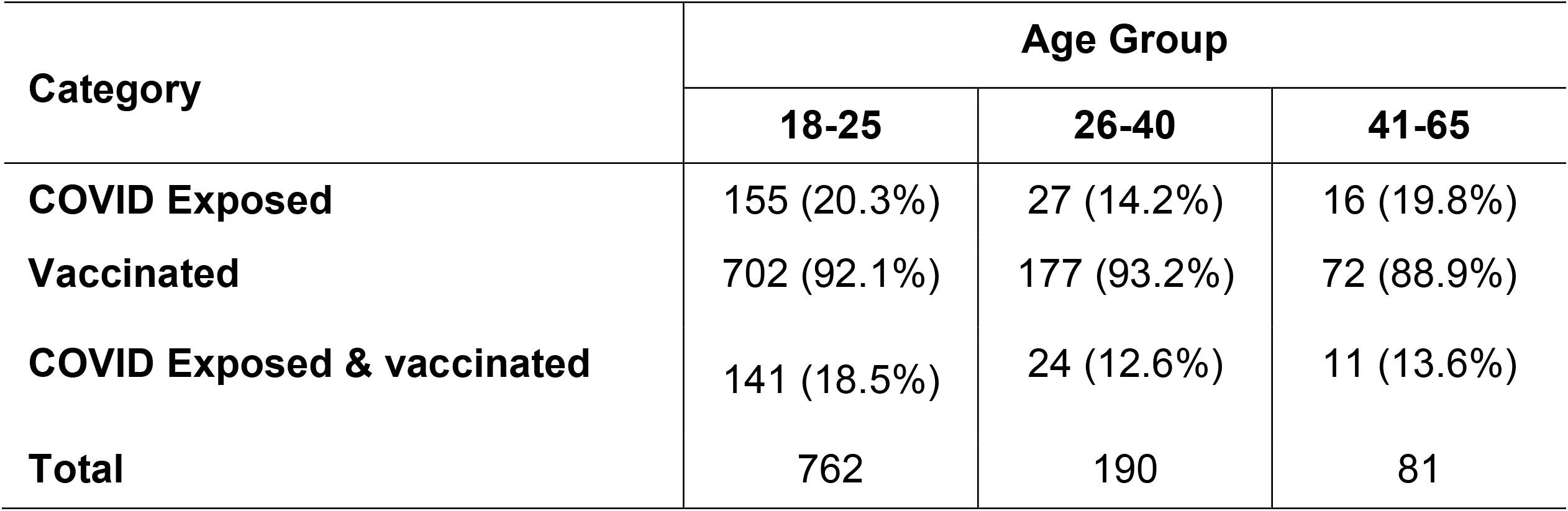
COVID and vaccine status by age group

### Seroprevalence

#### SARS-CoV-2 RBD of spike IgG and IgM antibodies

All serological assays were evaluated with the same set of 1064 serum samples (Table 4). Of 1064 individuals, the seroprevalence for anti-RBD IgG antibody was found to be 89.7% by Sienna-Clarity, 88.2% by Beckman, and 97% by MSD (Table 3). There were no significant differences in the seroprevalence of anti-RBD IgG antibodies cross the groups (all participants, students only, employee only, and randomly invited students).

**Table 3.** Anti-RBD and Anti-NC antibody seroprevalence status of the population

| Ab Sub-Type | Manufacturer | Antigen Detected | Name of the test | Assay type | Positives | Negative | Inconclusive |
| --- | --- | --- | --- | --- | --- | --- | --- |
| IgG | Beckman Coulter | RBD | Access SARS-CoV-2 IgG II (Semi-Quantitative) | Chemiluminescent Immunoassay (CLA) | 938 (88.2%) | 126 (11.8%) | 0 (0%) |
|  | Sienna-Clarity | RBD | Sienna-Clarity COVIBLOCK COVID-19 IgG/IgM Rapid Test | Lateral Flow (LFA) | 954 (89.7%) | 109 (10.2%) | 1 (0%) |
|  | MSD | RBD | V-PLEX COVID-19 Respiratory Panel 3 Kit | Electrochemiluminescent immunoassay | 1032 (97%) | 32 (3%) | 0 (0%) |
|  | Megna Health Inc. | Nucleocapsid | Rapid COVID-19 IgM/IgG Combo Test Kit | Lateral Flow (LFA) | 975 (91.6%) | 85 (8%) | 4 (0.4%) |
|  | MSD | Nucleocapsid | V-PLEX COVID-19 Respiratory Panel 3 Kit | Electrochemiluminescent immunoassay | 171 (16.1%) | 893 (83.9%) | 0 (0%) |
| IgM | Beckman Coulter | RBD | ACCESS SARS-CoV-2 IgM (Qualitative) | Chemiluminescent Immunoassay (CLA) | 87 (8.2%) | 977 (91.8%) | 0 (0%) |
|  | Salofa Oy | RBD | Sienna-Clarity COVIBLOCK COVID-19 IgG/IgM Rapid Test | Lateral Flow (LFA) | 8 (0.8%) | 1054 (99%) | 2 (0.2%) |
|  | Megna Health Inc. | Nucleocapsid | Rapid COVID-19 IgM/IgG Combo Test Kit | Lateral Flow (LFA) | 4 (0.4%) | 1055 (99.2%) | 5 (0.5%) |
| Total Ab (IgM, IgA, and IgG) | Bio-Rad | Nucleocapsid | Platelia SARS-CoV-2 Total Ab Assay | Enzyme-Linked Immunosorbent Assay (ELISA) | 210 (19.7%) | 841 (79%) | 13 (1.2%) |

\*Excluded Megna Health LFA results in further analysis due to higher rate of false positives

**Table 4.** Cohort characteristics and serological positive results by different assays

| Cohort |  |  | IgG (RBD from Spike Protein) |  |  | IgG (Nucleocapsid Protein) |  |
| --- | --- | --- | --- | --- | --- | --- | --- |
| Infection | Vaccine | n | CLIA<br>(Beckman) | LFA<br>(Sienna-Clarity ) | MSD<br>(MSD) | ELISA<br>(Bio-Rad) | MSD<br>(MSD) |
| YES | YES<sup>A</sup> | 180 | 177 (98.3%) | 179 (99.4%) | 179 (99.4%) | 101 (56.1%) | 69 (38.3%) |
|  | YES<sup>B</sup> | 2 | 2 (100%) | 2 (100%) | 2 (100%) | 2 (100%) | 2 (100%) |
|  | NO | 22 | 10 (45.5%) | 12 (54.5%) | 21 (95.4%) | 13 (59.1%) | 9 (40.9%) |
|  | NA | 1 | 1 (100%) | 1 (100%) | 1 (100%) | 1 (100%) | 1 (100%) |
| NO | YES<sup>A</sup> | 779 | 719 (92.3%) | 732 (94.0%) | 770 (98.8%) | 70 (8.9%) | 73 (9.4%) |
|  | YES<sup>B</sup> | 10 | 2 (20%) | 3 (30%) | 8 (80%) | 3 (30%) | 3 (30%) |
|  | NO | 60 | 21 (35%) | 19 (31.7%) | 41 (68.3%) | 20 (33.3%) | 14 (23.3%) |
|  | NA | 1 | 0 (0%) | 0 (0%) | 1 (100%) | 0 (0%) | 0 (0%) |
| NA | NA | 2 | 2 (100%) | 2 (100%) | 2 (100%) | 0 (0 %) | 0 (0%) |
| Total |  | 1064 | 938 (88.2%) | 954 (89.7%) | 1032 (97%) | 210 (19.7%) | 171 (16.1 %) |

<sup>A</sup>Pfizer/BioNTech, Moderna, Janssen, AstraZeneca, Covishield; <sup>B</sup>Sinopharm, Sinovac, Covaxin

Among 182 participants who self-reported COVID infection and were vaccinated, 179 (98.4%) tested positive by Beckman, 181 (99.5%) by Sienna-Clarity LFA, and 181 (99.5%) by MSD for anti-RBD antibody. Among 22 participants who self-reported COVID infection and were not vaccinated, 10 (45.5%) tested positive by Beckman immunoassay, 12 (54.5%) by Sienna-Clarity, and 21 (95.4%) by MSD for anti-RBD antibody. Among 789 participants who self-reported no-COVID infection and were vaccinated, 721 (91.4%) tested positive by Beckman, 735 (93.2%) by Sienna-Clarity, and 778 (98.6%) by MSD for anti-RBD antibody (Table 4).

#### SARS-CoV-2 NC antibodies

Overall, the seroprevalence for total anti-NC was 19.7% by Bio-Rad and 16.1% for anti-NC IgG by MSD, but 91.6 % by Megna. We excluded results from the latter due to high false-positive results (91.6%) (Table 3) since the seroprevalence was estimated at 34.2% in September 2021 from the nationwide commercial lab in Arizona based on the CDC website ^4^.

Among 205 participants who self-reported COVID infection regardless of vaccination status, 117 (57.1%) by Bio-Rad and 81 (39.5%) by MSD tested positive for anti-NC antibody levels (Table 4). Interestingly, almost 80% (n=840) of the participants reported no known history of infection regardless of vaccine status (excluding 10 participants who received attenuated parasite vaccines*^B^). However, 10.7% (n=20+70=90) and 10.4% (n=73+14=87) tested positive for anti-nucleocapsid antibodies by the ELISA and the MSD assay without recalling at least one SAS-CoV-2 infection (Table 4), presumably representing occult infections.

#### Comparison of assays performances

The Venn diagrams show the overlapping distribution of positive results for each assay. For seropositive responses to the RBD of the spike protein, 926 specimens were positive by all three of Sienna-Clarity, Beckman, and MSD, whereas 75 specimens were positive only by MSD (Figure 2A). Based on the same sample population, the percentage of positive results for all three assays for anti-RBD IgG were comparable (90%, 88%, and 97% respectively; Figure 2). However, only Beckman and MSD provided the antibody levels provided a quantitative number which allowed us to track antibody levels post vaccination and monitor how long immunity persisted. In addition, Figure 3A showed the correlation of the values of anti-RBD IgG between two assays. The anti-RBD antibody results by Beckman correlated strongly with the results by MSD (r=0.79).

**Figure 2:**
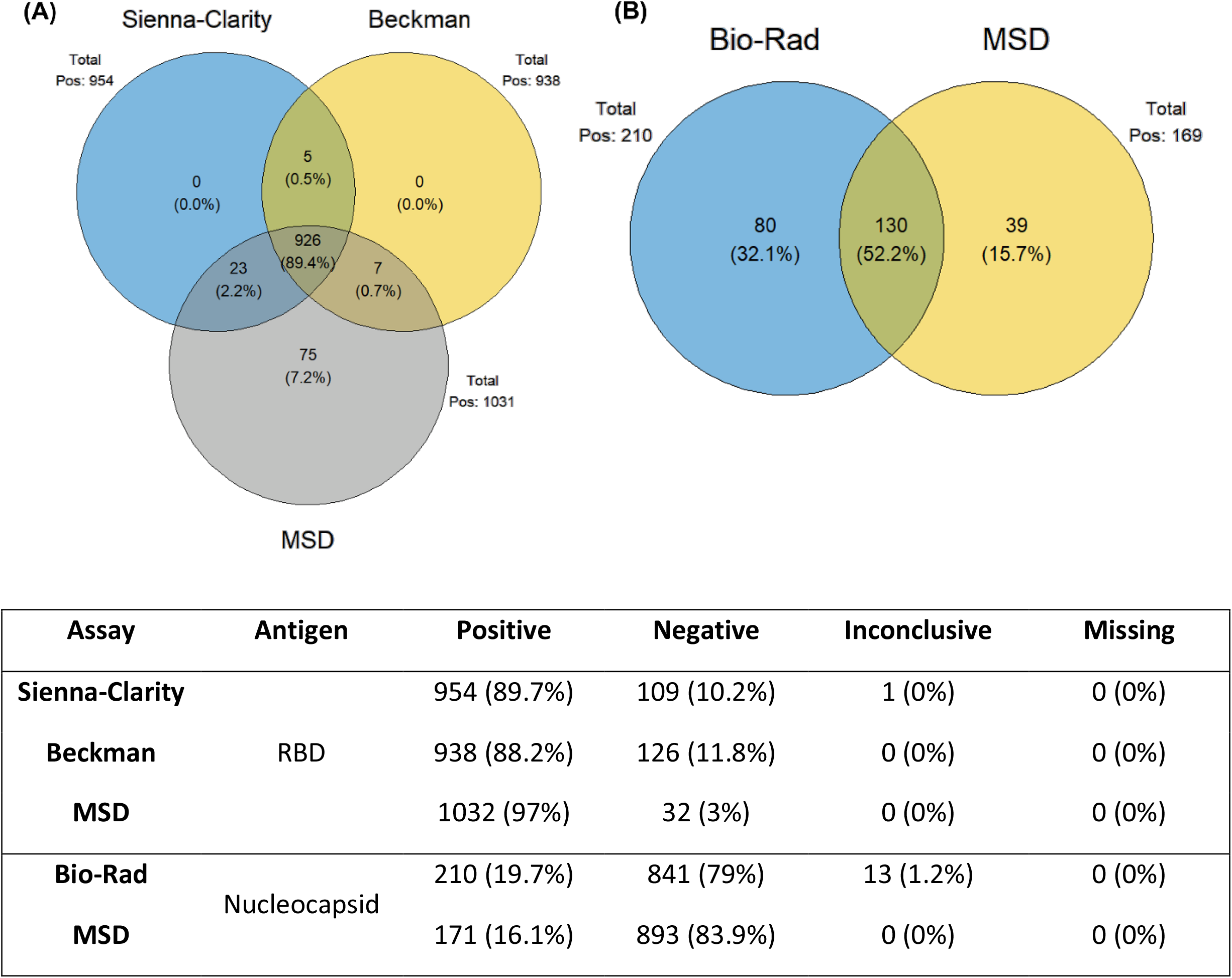
Comparison of assay performances. Venn diagrams showing overlap of positive results of (A) RBD of Spike and (B) Nucleocapsid from different assays.

**Figure 3:**
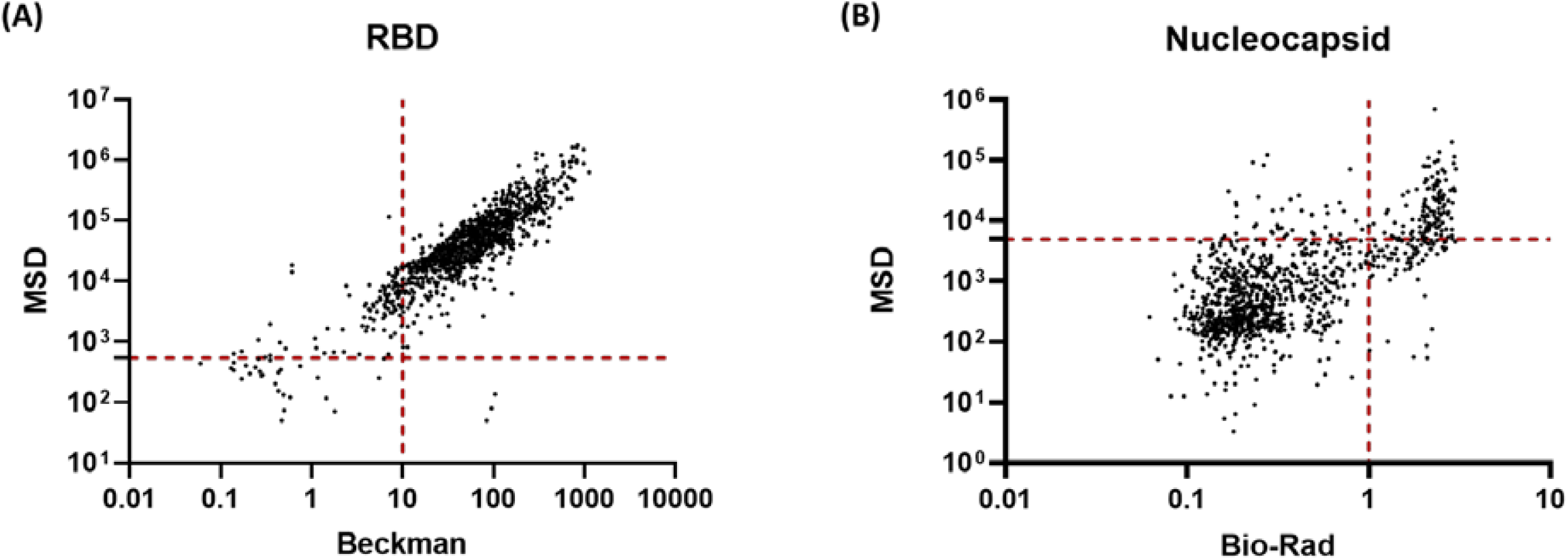
Correlations between antibody results by different assays (A) Correlation between the value of anti-RBD IgG by Beckman and the MSD assay. (B) Correlation between the value of total anti-NC by Bio-Rad assay and the value of anti-NC IgG by the MSD assay. A red dotted line indicated the cut-off line. All test values equal to or greater than this line is considered positive.

For seropositive NC, 130 specimens were positive by both Bio-Rad and MSD, whereas 80 specimens were positive only by Bio-Rad and 39 specimens by MSD (Figure 2B). The correlation of the values of anti-NC antibody level between Bio-Rad and MSD was weak (r=0.34) (Figure 3B).

#### Anti-RBD IgG antibody levels after vaccination

Anti-SARS-CoV-2 antibody persistence in the first six months after COVID vaccination decreased over time ^19-21^. Here, we examined the relationship between the anti-RBD antibody titers of participants who received COVID vaccines and the number of days after vaccination using linear regression and summarized in Figure 4. As indicated in Figure 4, antibody titers varied widely, but there was clear trend towards lower titers over time. All vaccines have the same trend; we only report Modern and Pfizer in Figure 4; the other vaccines are reported in supplementary Figure 1. Participants who received 2 doses of Moderna vs. Pfizer trended towards higher antibody titers, which lasted longer, although these results were not statistically separable.

**Figure 4:**
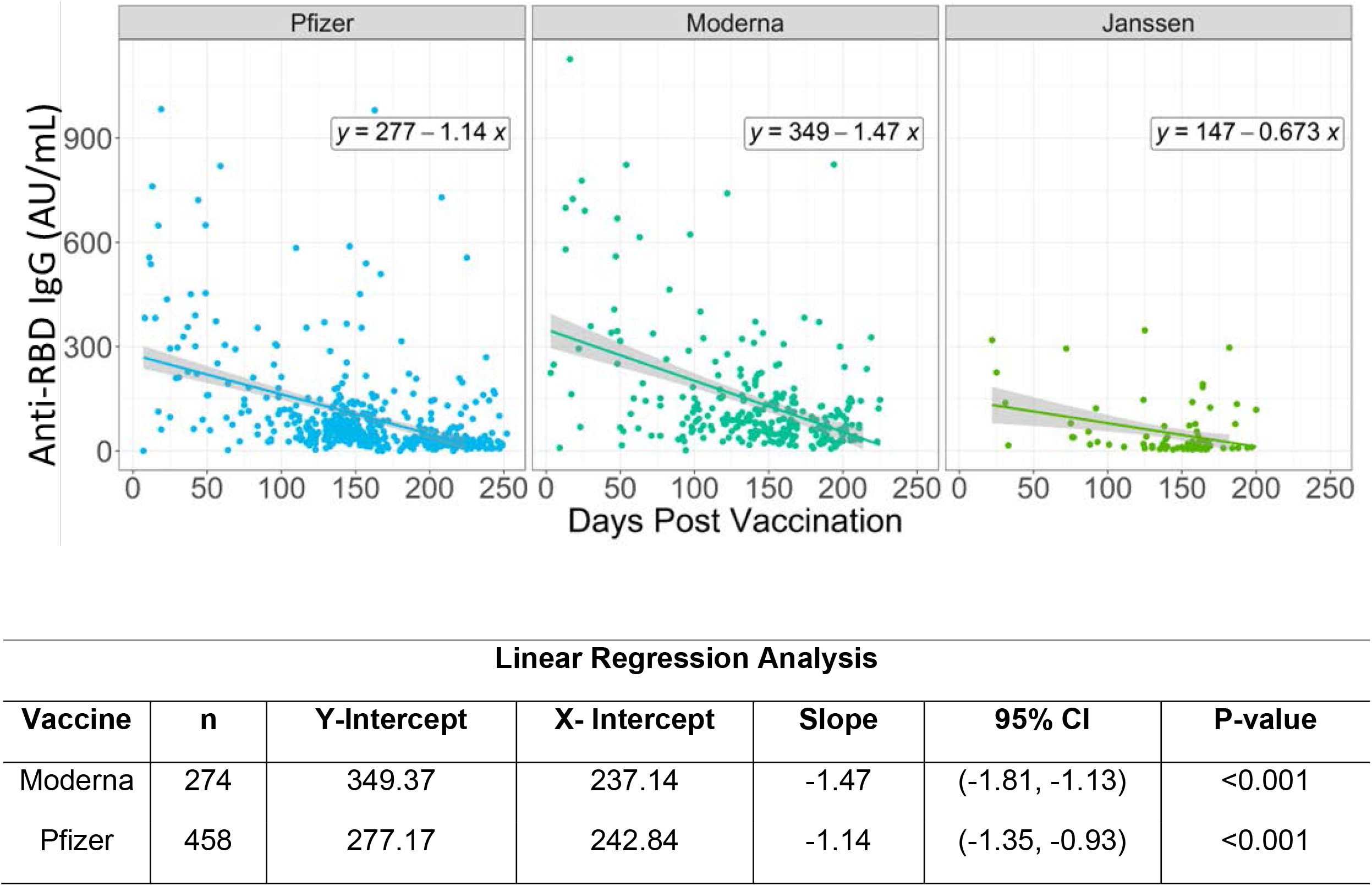
Anti-RBD antibody decay post-vaccination. The antibody level is determined by the Beckman immunoassay. The linear regression of different vaccines to estimate vaccine decay.

#### RBD Antibody responses following vaccination/infection

Participants were first classified into different groups based on their vaccine and infection status (vaccine only, previous COVID infection only, and both) and then further categorized them based on the time between their most recent vaccination/infection date and the collection date (0-3, 4-6, ≥7 months). In each group, the median level of anti-RBD antibody levels was higher in the subgroups of vaccinated participants with COVID infection than those with vaccination or infection only. In every group, the lowest median anti-RBD antibody level was detected in the participants who were never vaccinated. There were no samples in the group of participants with infection after 4-6 months. Although anti-RBD antibody levels declined over time for all groups, median antibody levels in both vaccinated and infected or vaccination-only groups remained above the cut-off 7 months after either infection and/or vaccination, whereas median antibody levels in the infection only group dropped below the cut-off by 7 months post infection (Figure 5).

**Figure 5:**
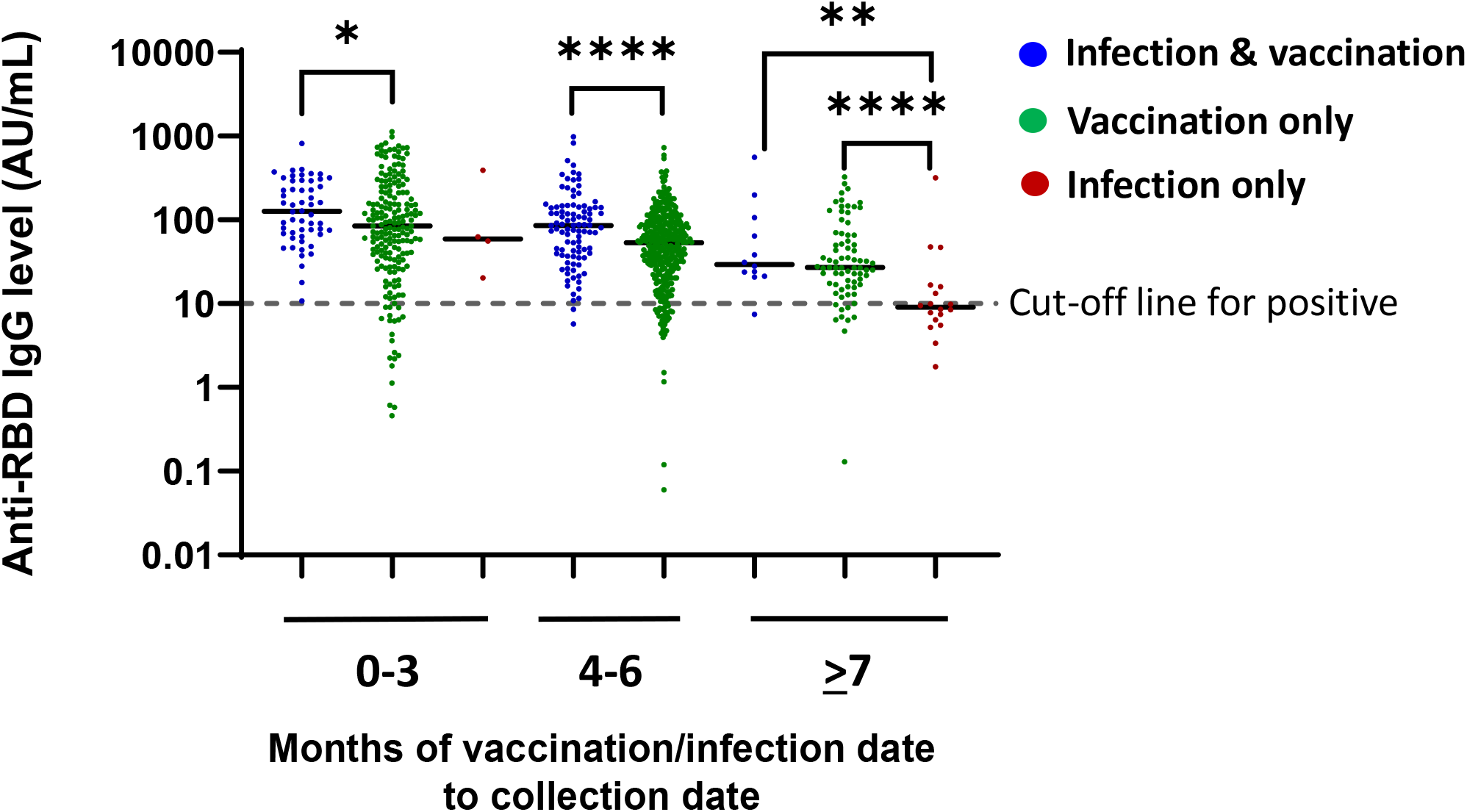
Anti-RBD antibodies in participants who had previous COVID infection or COVID vaccines or both. Participants were categorized by the vaccine or COVID infection they got and the number of months between their vaccination/infection and their blood sample collection. Anti-RBD IgG level is measured by Beckman immunoassay. Cut-off defined per manufacturer. *P value is calculated by the Mann-Whitney test

#### Increased anti-RBD IgG levels after breakthrough infection

Next, we investigated whether breakthrough COVID was associated with improved immune response. Participants were classified into three groups (breakthrough infection, hybrid immunity which is the participant who received vaccination after SARS-CoV-2 infection, and vaccine only). We had 645 fully vaccinated individuals, 19 individuals with 2 doses of vaccine after COVID infection (hybrid immunity), and 12 fully vaccinated individuals with breakthrough infection. Anti-RBD IgG values were significantly increased in both breakthrough and hybrid immune groups compared to vaccine only. In addition, the breakthrough infection group had significantly higher antibody levels compared to the hybrid immunity group, showing an association between breakthrough and enhanced immune response (Figure 6).

**Figure 6:**
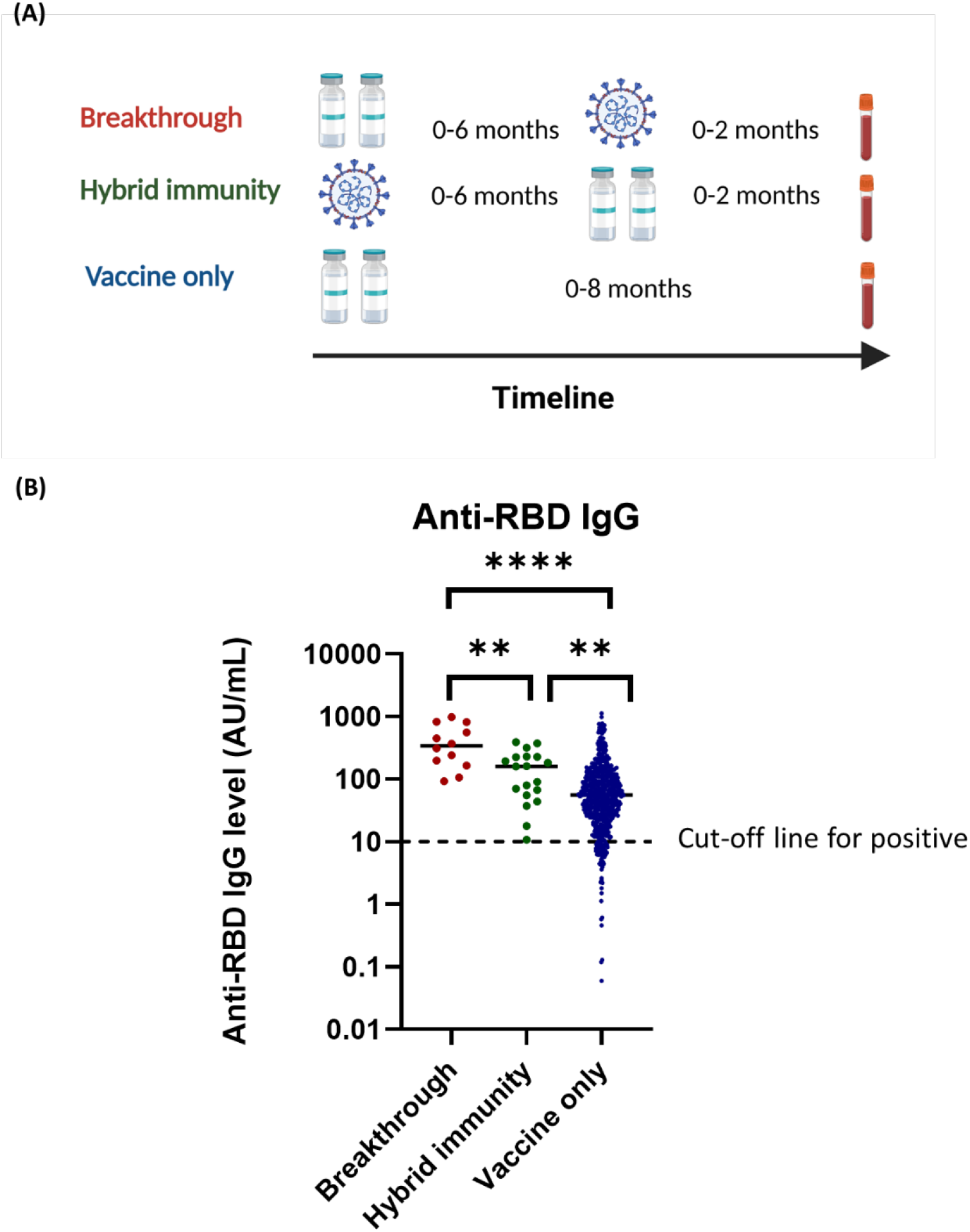
Anti-RBD antibodies after breakthrough infection, hybrid immunity, and vaccine only. (A) Participants were categorized based on the order and approximate time scale of COVID infection and vaccination for each group. The blue bottle indicates a dose of vaccine, the virus indicates natural infection with SARS-CoV-2 based on the participant’s self-reported, and the red vial indicates blood collection. (B) Anti-RBD IgG level is measured by Beckman immunoassay *P value is calculated by the Mann-Whitney test

#### Increased anti-NC IgG antibody levels after infection

We also analyzed the antibody response to the SARS-CoV-2 NC protein in previously infected participants. However, the EUA-approved ELISA for NC antibodies that we used (Bio-Rad) is only a qualitative assay. To understand the trend of NC antibody levels post-infection, the MSD (Meso Scale Discovery) immunoassay from Meso Scale Diagnostics was applied, which uses ELISA-based quantitative detection. This assay was already verified with clinical samples, even though it is not an EUA-approved assay. Participants were categorized into different groups (0-1, 2-3, 4-5, 6-7, 8-9, 10-11, 12-13, 14-15, >15 months) depending on the interval between their infection date and collection date. Like RBD antibody levels, the NC antibody levels decreased over time, dropping somewhat faster than the ant-RBD antibodies in these data (Figure 7 & Supplementary Figure 2).

**Figure 7:**
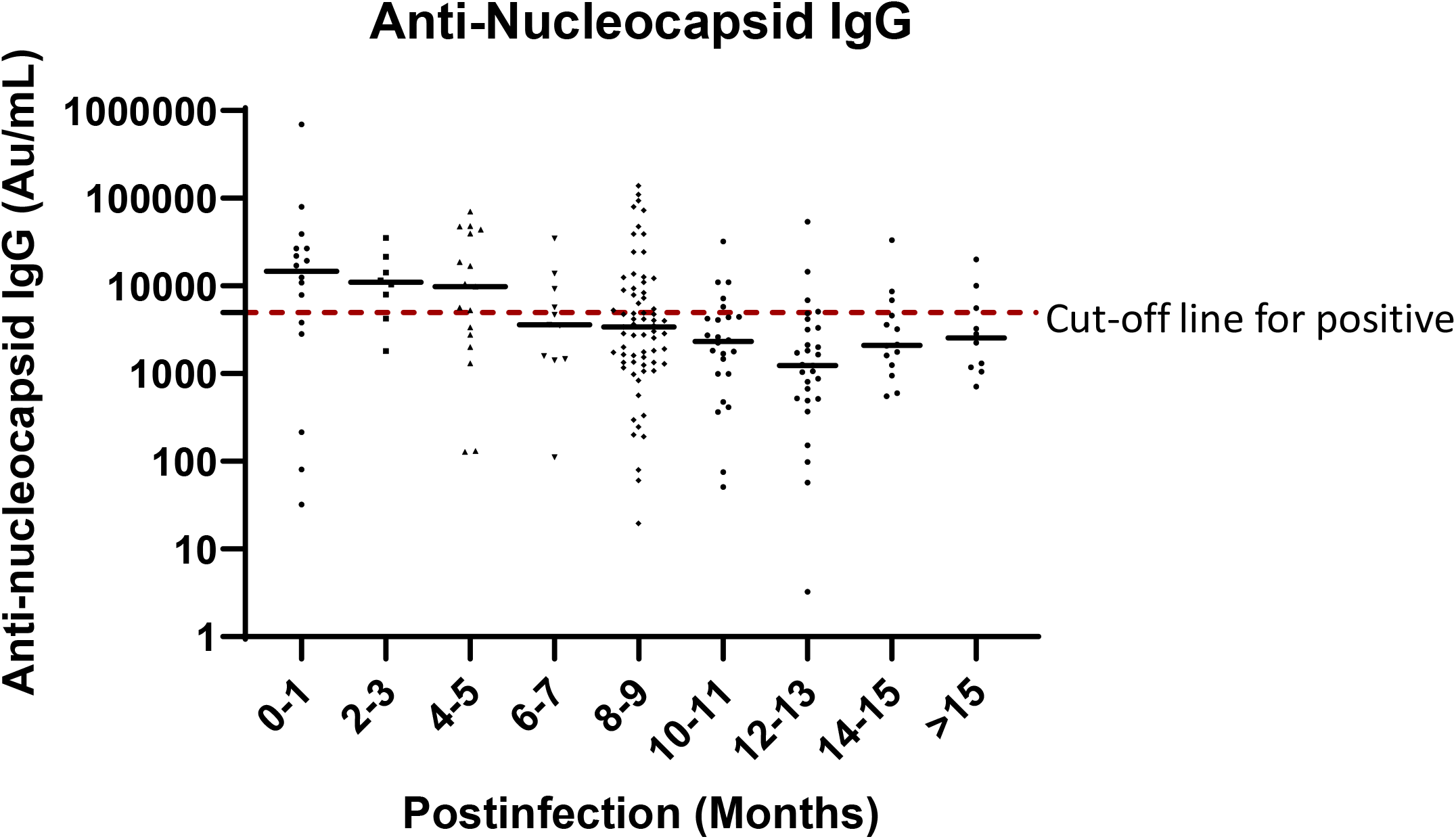
Anti-nucleocapsid antibodies after COVID infection. Participants were categorized by the COVID infection they got and the number of months between their infection and their blood sample collection. Anti- nucleocapsid IgG levels were measured by ELISA. *P value is calculated by the Mann-Whitney test

## Discussion

In the fall of 2021, over 79,000 students returned to campus for in-person classes coincident with a large increase in COVID incidence during the Delta wave in Maricopa County, AZ. We observed only 0.38% (4 positives out of 1064) active COVID positivity based on saliva qPCR on the day of sample collection from the serosurvey study in the ASU community. Notably, those with symptoms were asked not to participate.

### Vaccination compliance among the participants was very high

In the ASU community, 92% of participants have self-reported to had at least one dose of a COVID-vaccine. By comparison, only 85% of college students in the U.S. enrolled in spring or fall 2022 were vaccinated based on a nationally representative survey by the American College Health Association ^22^. We believe, AUS’s proactive communications to parents and students helped with increased rate of vaccination. Most of the vaccinated participants at ASU received the Moderna and Pfizer mRNA vaccines that have shown great effectiveness after the second dose. However, as previously noted, the antibodies produced by Pfizer’s COVID-19 vaccine decline faster than those produced by the Moderna vaccine after 6 months of vaccination ^23^. We observed a similar trend in our study. Based on anti-RBD antibodies levels from Beckman it showed that a higher anti-RBD IgG antibody level lasted longer in the participants who received 2 doses of Moderna compared to those who received 2 doses of Pfizer. This is probably due to the higher amount of RNA in Moderna (Figure 4).

Interestingly, 7 out of 978 participants who self-reported having received a COVID vaccine, tested negative for anti-RBD antibodies by all three assays in our study. Three out of 7 participants were vaccinated for more than 5 months (165, 168, and 216 days) with Pfizer vaccines leading to potential antibody decay based on figure 4. One out of 7 participants only received Pfizer for 7 days and antibody was likely not generated. It is known that there is substantial variation between individuals in the immune response to vaccination ^24^. Other two out of 7 participants received Covaxin for 56 and 80 days and the level of anti-RBD antibodies in Covaxin was significantly lower than other vaccines. Another one out of 7 participants received AstraZeneca for more than 3 months (101 days), showing that Anti-RBD antibody levels from AstraZenecca started to wane after 2 months (Supplementary Figure 1) which was similar to previously reported result from other group ^25^.

### The participants were tested negative for NC antibody after 6 months of post infection with COVID

In the ASU community, 19.3% of participants (n=205) self-reported they had previous COVID infection; however, we only found 57% (n=117/205) and 39.5% (n=81/205) of participants from this group tested positive for NC antibody by Bio-Rad and MSD, respectively (Table 5). This could be due to antibody decay since their SARS-CoV-2 exposure. The median NC antibody levels fell below the positivity cutoff 6 months after infection, based on our MSD data (Figure 7). Also, by 8 months post infection, 50% of participants from this group had undetectable NC antibodies. This finding was common with other serological studies, where the NC antibodies started to decline after a few months post infection and half the of participants have undetectable NC antibodies by 8 months post infection ^26,27^.

Among 847 self-reported no previous COVID infection participants (excluding 10 participants who received attenuated parasite vaccines*^B^), 10.6% (n=90) and 10.3% (n=87) tested positive for anti-NC antibody by Bio-Rad and MSD which means these 10% of participants had a COVID infection in the past without realizing it (Table 4). It could be these participants had mild or asymptotic previous COVID-19 infections.

### SARS-CoV-2 antibodies and breakthrough infections

A main finding of this serological survey was that the participants who had breakthrough infection had higher anti-RBD IgG compared to those who were fully-vaccinated and also had prior infection (Figure 6), which agrees with previous studies ^28,29^. Considering that the antibody developed by B cells multiply after each exposure through infection or vaccination, these results were expected. First, the highest anti-RBD antibody levels were in the combined vaccination and infection group and most likely represent an accumulation of antibodies produced after each exposure. Second, the anti-RBD antibody level in the infection only group decayed faster than the participants who received vaccines only. The participants here predominantly received the Pfizer and Moderna vaccines, which may be particularly efficient at evoking a durable anti-RBD response. Similar observations were made by Dashdor et al, that participants who received the Sinopharm vaccine (whole virus) had lower antibody levels compared to Pfizer/Moderna vaccine (spike protein) ^30^.

### Limitations

Study limitations need to be noted in this study, including samples with an unknown degree of selection bias due convenience sample, self-reported COVID test results, and vaccine status. Participants were only tested once for antibody, thus lacking longitudinal data to compare antibody waning rates in individuals. In addition, the number of breakthrough infections and infection only were relatively small. Conducting longitudinal studies at university settings will provide valuable information about vaccine efficacies, infection spread among vaccinated individuals and provide mitigation regarding policies that work when implemented appropriately, during current and future pandemics.

## Data Availability

All data produced in the present work are contained in the manuscript

## Acknowledgements

We would like to thank ASU EHS team for their logistical support and IBC oversight, communication team for their support in communicating study materials to ASU community, members of our clinical coordination team Jerome Woodfin, Will Taylor and Abriana Gonzales for their help in sample accessioning, and Scarlett Goins-Heisler, Harrison Bell, Ankita George, Andrew Garcia, Guillermo Trivino, and David Rainford for their help in processing the blood tubes during the day of the serosurvey. We thank Steven Winn, and Jessica Lukosus for their help in managing the event and procurement of supplies. Funding provided by Arizona State University Knowledge Enterprise.

