## Supplemental tables and figures for "Serosurvey of SARS-COV-2 at a large public university"

**Supplementary Table 1:** Manufacturer reported sensitivity and specificity of EUA authorized tests used in this study

| Assay | Sensitivity (positive percent agreement at >/= 15 days post symptom onset) | Specificity |
|---|---|---|
| Chemiluminescent assay: Beckman (Anti-RBD IgG) | 98.9% | 100% |
| Later flow assay: Clarity (Anti-RBD IgG) | 96.15% | 100% |
| Later flow assay: Megna (Anti-NC IgG) | 95% | 99.3% |
| ELISA: Bio-Rad (Anti-NC IgM/IgG/IgA) | 100% | 98.86% |
| Electrochemiluminescence assay: MSD* (Anti-NC IgG) | 93.8% | 100% |
| Electrochemiluminescence assay: MSD* (Anti-RBD IgG) | 98.3% | 98.5% |

*****MSD is not a EUA authorized test. It is a validated assay meets the clinical laboratory standards institute guidelines.

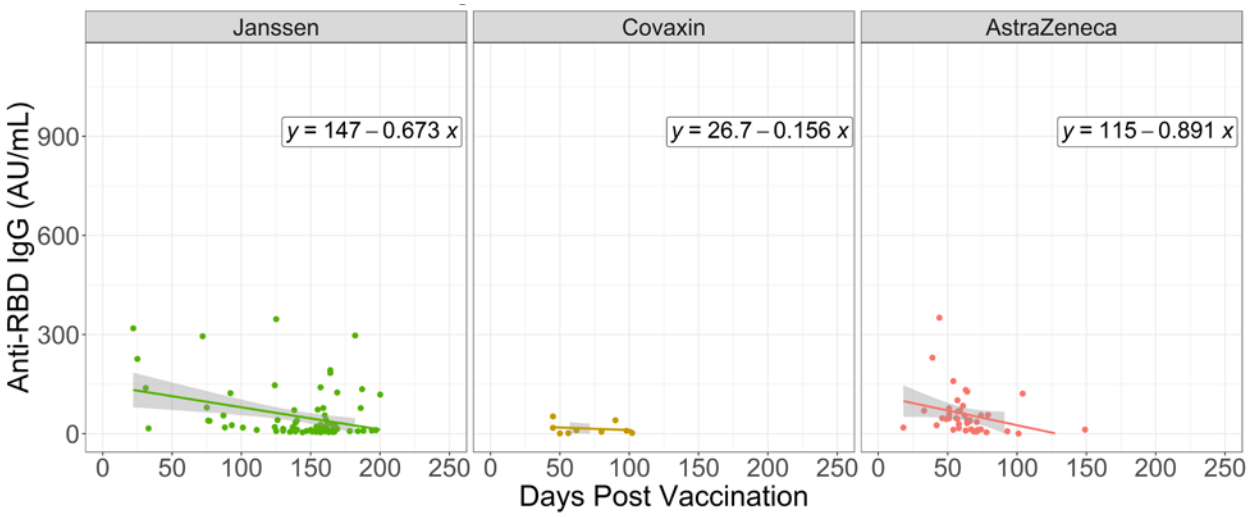


**Supplementary Figure 1**: **Anti-RBD antibody decay post-vaccination.** The antibody level is determined by the Beckman immunoassay. The linear regression of different vaccines to estimate vaccine decay.

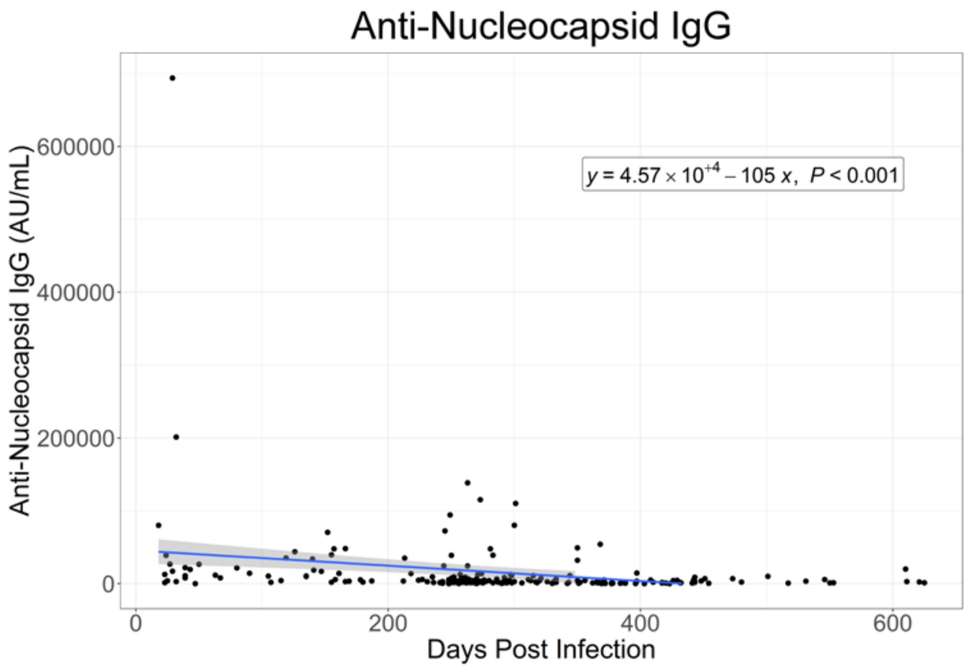


**Supplementary Figure 2**: **Anti-Nucleocapsid antibody decay post-infection.** The antibody level is determined by the MSD. The linear regression was to estimate antibody decay.
